# Evaluating the causal effect of mitochondrial dysfunction on Alzheimer’s and Parkinson’s disease using Polygenic Risk Scores and Mendelian Randomization

**DOI:** 10.1101/2025.09.25.25336251

**Authors:** Aadrita Chatterjee, Brian Alvarez, Rakshya U Sharma, Caroline Jonson, Heather M. Wilkins, Judy Pa, Russell H. Swerdlow, Alison Goate, Kristine Yaffe, Shea J Andrews, the Alzheimer’s Disease Genetics Consortium and the Alzheimer’s Disease Neuroimaging Initiative

**Affiliations:** Department of Psychiatry and Behavioral Sciences, University of California San Francisco, 505 Parnassus Ave, San Francisco, CA, USA Department of Psychiatry and Behavioral Sciences, University of California San Francisco, 505 Parnassus Ave, San Francisco, CA, USA; Department of Genetics and Genomic Sciences, Icahn School of Medicine at Mount Sinai, New York, NY, USA; Center for Alzheimer’s and Related Dementias, National Institutes of Health, Bethesda, MD, USA; DataTecnica LLC, Washington DC, USA; Memory and Aging Center, Department of Neurology, Weill Institute for Neurosciences, University of California, San Francisco, San Francisco, CA, USA; Department of Neurology, University of Kansas Medical Center, Kansas City, KS 66160, USA; Department of Biochemistry and Molecular Biology, University of Kansas Medical Center, Kansas City, KS 66160, USA; University of Kansas Alzheimer’s Disease Research Center, University of Kansas Medical Center, Kansas City, KS, USA; Alzheimer’s Disease Cooperative Study (ADCS), Department of Neurosciences, University of California, San Diego, CA, USA; Department of Epidemiology and Biostatistics, University of California, San Francisco, San Francisco, CA, USA

**Keywords:** Mitochondrial DNA copy number, Alzheimer’s disease, Parkinson’s disease, Mendelian Randomization, Polygenic Risk Scores, genetic correlations

## Abstract

**INTRODUCTION:** Mitochondrial DNA copy number (mtDNAcn), a measure of mitochondrial genomes per nucleated cell, has an unclear causal relationship with AD and PD. We integrate genetic correlation, Polygenic Risk Scores (PRS), and Mendelian Randomization (MR) to assess whether mtDNAcn influences the risk of AD and PD, and evaluate how study-specific factors in mtDNAcn genome-wide association studies (GWAS) may distort these causal estimates.

**METHODS:** Using GWAS of four mtDNAcn measures, AD, AD/dementia, and PD, we evaluated genetic correlations, generated ancestry-normalized PRS in the AD Genetics Consortium (N=27,383), and applied MR methods including Latent Heritable Confounder MR (LHC-MR).

**RESULTS:** Across the four mtDNAcn GWAS, only one was consistently associated with AD/dementia and PD, with genetic correlations and PRS analysis showing negative correlations and MR indicating that higher mtDNAcn reduced AD/dementia and PD risk.

**DISCUSSION:** Higher blood-based mtDNAcn was causally associated with reduced risk of AD/dementia and PD, with limited evidence to suggest a bidirectional effect.

**Research In Context:** *Systematic Review:* Mitochondrial dysfunction, measured by mitochondrial DNA copy number (mtDNAcn), has been linked to Alzheimer’s disease (AD) and Parkinson’s disease (PD). However, Mendelian randomization (MR) studies on this relationship have shown inconsistent results, have not applied advanced MR methods that address prior limitations, or examined study-specific biases.

*Interpretation:* Using genetic correlations, polygenic scores, and Mendelian Randomization, we triangulated evidence across complementary methods. We found that results varied depending on the dataset (e.g., clinically diagnosed AD vs. family history of AD) and study design factors such as mtDNAcn measurement techniques. Despite these biases, higher mtDNAcn was consistently associated with a lower risk of AD and PD, supporting a mitochondrial mechanism in both diseases.

*Future directions:* Our findings highlight mtDNAcn as a potential biomarker for AD/PD, emphasizing the importance of measurement methods. Future research is needed to explore the biological pathways underlying this relationship.

**Highlights:**

- Genetically predicted higher mtDNAcn is causally associated with lower AD and PD risk
- AD genetic liability is causally associated with higher mtDNAcn, possibly as a compensatory response
- mtDNAcn is a potential early biomarker of mitochondrial dysfunction in AD/PD

## 1. Background

Mitochondria are membrane-bound organelles that play a central role in essential cellular processes, including managing oxidative stress, beta-oxidation of fatty acids for energy production, and facilitating programmed cell death^1^. These processes work together to maintain mitochondrial integrity and ensure efficient cellular function by removing or replacing damaged or dysfunctional mitochondria^1^. An important marker of mitochondrial health is mitochondrial DNA copy number (mtDNAcn), which refers to the number of mitochondrial genomes present per nucleated cell^2^. This measure has been widely used as a proxy for mitochondrial function and is implicated in various physiological and pathological conditions, including aging, metabolic disorders, and neurodegenerative diseases^2^. The current gold standard for measuring mtDNAcn is quantitative PCR (qPCR); however, mtDNAcn can also be estimated using microarray probe intensities and whole-genome sequencing (WGS).^3^

Mitochondrial dysfunction plays a prominent role in two of the most common neurodegenerative diseases - Alzheimer’s disease (AD) and Parkinson’s disease (PD)^4,5^. Higher mtDNAcn has predominantly been associated with reduced risk of AD and PD, better cognitive function, and lower tau pathology^6–8^. Confounders, such as cell type heterogeneity, and reverse causation, complicate the interpretation of associations between mtDNAcn and neurodegenerative disorders. Different cell types contain varying levels of mitochondrial DNA; for instance, platelets, due to their high energy demands, contain significantly more mitochondrial DNA than other cells but lack nuclear DNA^[2]2^. Additionally, elevated mtDNAcn may reflect compensatory upregulation—an adaptive response to impaired mitochondrial function—raising the question of whether mtDNAcn is a cause or consequence of disease^2^.

Mendelian randomization (MR) studies use genetic instruments to determine the causal effect of an exposure on an outcome, and they are less susceptible to confounding and reverse causation. MR studies examining the causal effect of mtDNAcn on AD and PD have reported conflicting findings. While some two-sample MR analyses suggest that higher blood mtDNAcn is causally associated with reduced AD risk, other studies have found no strong evidence of a causal effect on either AD or PD^5,7,9,10^. Furthermore, these analyses often do not account for unmeasured confounding and typically rely on a limited set of genetic variants.

In this study, we integrate genetic correlation, PRS, and multiple MR frameworks to assess whether genetically predicted blood mtDNAcn influences the risk of AD and PD, and to evaluate how study-specific factors in mtDNAcn GWAS may distort these causal estimates. Genetic correlations test whether there is shared genetic architecture between traits due to pleiotropy^11^. PRS test whether genetic predisposition to one particular trait, such as mtDNAcn, is associated with a second trait ^12^. However, while genetic correlations and PRS establishes correlation, they do not infer causality.

To assess the causal direction of the effect of mtDNAcn on AD/PD, we can use MR, applying state-of-the-art approaches such as latent heritable confounder MR (LHC-MR) to account for unmeasured confounding and obtain unbiased causal estimates^13,14^. Furthermore, since not all mtDNAcn GWAS adjust for platelet count—a major biological confounder—we also implement Multivariable Mendelian Randomization (MVMR) to estimate both direct and indirect effects of each exposure on the outcome^15^. Finally, by leveraging multiple studies that quantify mtDNAcn using different methods, we can assess the robustness of our findings and determine whether the observed causal effects reflect true biological relationships or are influenced by study-specific measurement biases. Together, these approaches allow us to test both association and causation while addressing key methodological limitations in prior work.

## 2. Methods

We employed a combination of analytical approaches to investigate whether mtDNAcn plays a causal role in AD and PD. Genetic correlations and PRS were used to determine whether mtDNAcn has a shared genetic architecture with AD and PD, establishing a basis for causal inference. Forward direction MR analyses were used to evaluate whether genetically determined mtDNAcn is causally associated with AD or PD. Lastly, we applied MVMR to adjust for platelets and LHC-MR to provide more robust estimates and confirm univariate MR analyses. An overview of the study design is provided in Supplementary Figure 1.

### 2.1 Data Sources

We used GWAS summary statistics for each exposure and outcome dataset (Table 1 and Supplementary Table 1). The first GWAS used for AD involved clinically diagnosed AD cases (AD)(n= 94,437, n_loci_ = 25 LOAD risk loci)^16^. The second GWAS used clinical case-control status and family history of dementia to perform a GWAS-by-proxy (GWAX) (n = 788,989, n_loci_ = 72) of AD and related dementias (AD/dementia)^17^. The GWAS for PD (n ∼1.4 million, n_loci_ = 90) used 37.7K cases, 18.6K UK Biobank proxy-cases (having a first-degree relative with PD), and 1.4M controls^18^.

**Table 1.**
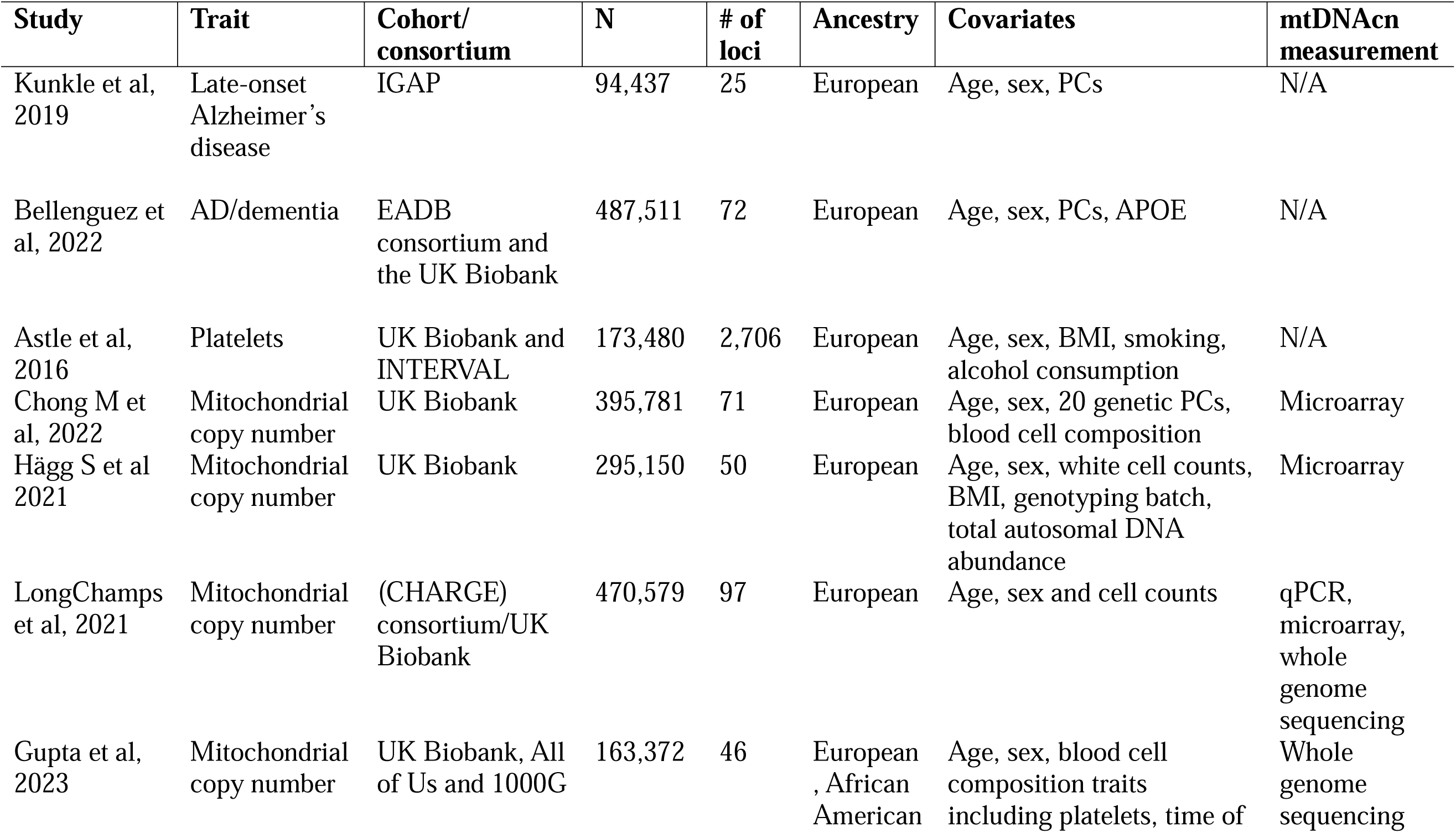
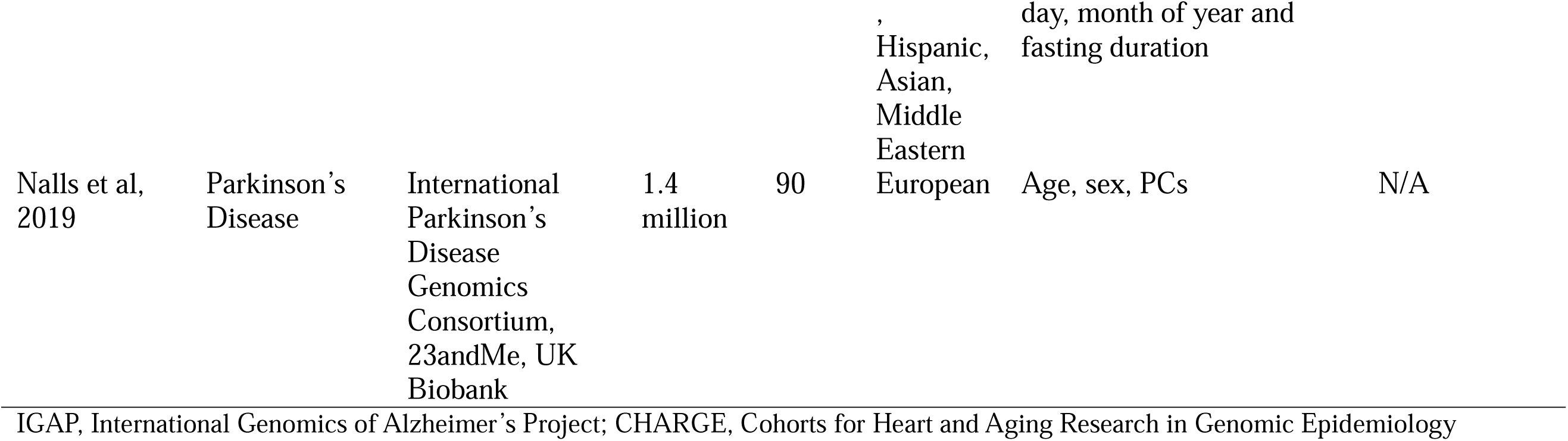
Description of GWAS datasets used in this study.

Blood-derived mtDNAcn came from four different GWASs. The Longchamps et al (n= 470,579, n_loci_ = 97) quantified mtDNAcn through WGS, quantitative PCR (qPCR), and microarray probe intensities^19^. Hägg et al (n= 295,150, n_loci_ = 50) estimated mtDNAcn by the weighted intensities of genotyping probes^20^. Chong et al (n= 395,781, n_loci_ = 71) measured mtDNAcn by utilizing a microarray-based method called Automatic Mitochondrial Copy Number (AutoMitoC)^21^. This was unique in the following ways (1) replacement of autosomal signal normalization of common variants with globally rare variants, (2) assessment of the association of corrected probe signal intensities with off-target genome intensities to detect cross-hybridizing probes, and (3) MT signal is ascertained via principal component analysis, rather than by median signal intensity. Finally, Gupta et al (n=163,372, n_loci_ = 46) quantified mtDNAcn using WGS^22^.

To account for platelet abundance in MVMR, we used a GWAS for platelets (n=173,480, n_loci_ = 2,706).

All datasets included age and sex as covariates and were standardized using the gwas2vcf pipeline^23^. Supplementary Table 1 details the study design, eligibility and quality control measures.

### 2.2 Genetic Correlations

We formatted and munged GWAS using Linkage Disequilibrium Score Regression (LDSC) version 1.0 and estimated genetic correlations between each trait using GeNetic cOVariance Analyzer (GNOVA)^24,25^. GNOVA is a principled framework that leverages GWAS summary statistics to assess annotation-stratified genetic covariance between traits^25^. Genetic correlations allow us to see the shared genetic basis of our traits of interest and can result from horizontal or vertical pleiotropy. They can be quantified as strong (rg>0.6), medium (rg 0.2 - 0.6) or weak (rg<0.2)^11^.

### 2.3 Polygenic Risk Scores

PRS estimate an individual’s genetic liability to a trait or disease by aggregating the effects of multiple single nucleotide polymorphisms (SNPs), each weighted by its effect size derived from GWAS^12^. We constructed mtDNAcn PRS using data from the Alzheimer’s Disease Genetics Consortium (ADGC), one of the largest multi-ancestry AD datasets. Genotyping and imputation procedures for the contributing cohorts have been described previously^26^. For this study, we applied genotype quality control to TOPMed-imputed datasets using GenoTools^27^. Variants were filtered to exclude those with a call rate < 95% and minor allele frequency < 1%. Samples were evaluated for relatedness (kinship coefficient > 0.0884) and excessive heterozygosity. Related individuals and potential duplicate samples (IBS > 0.354) were identified and removed, leaving one representative sample per pair.

mtDNAcn summary statistics were filtered to HapMap3 SNPs, and score files were generated using PRS-CS-auto (v1.1.0) to automatically learn the global shrinkage parameter (□) using a fully Bayesian approach^28^. PRS were calculated and applied to the ADGC using PGSC-calc (v2.0.1) for ancestry normalization^29^. Genotypes were lifted over to the GRCh38 reference genome and matched to the ADGC dataset using our custom score file^29^. We used the Human Genome Diversity Project + 1000 Genomes Project reference panel for ancestry inference and excluded the APOE e4 region (19:44408822-19:45408822, build 38) due to confounding^29^. To determine the association of PRS with AD status in ADGC, we performed logistic regression adjusting for age, sex, *APOE*-ε4 status, and population stratification (PC 1-4).

To validate that our PRS models predicted mtDNAcn, we evaluated the association of each mtDNAcn PRS with mtDNAcn in participants contributing to ADGC from the Alzheimer’s Disease Neuroimaging Initiative (ADNI) database (adni.loni.usc.edu) (Table 2 and Supplementary Table 2). ADNI was launched in 2003 as a public-private partnership, led by Principal Investigator Michael W. Weiner, MD. The primary goal of ADNI has been to test whether serial magnetic resonance imaging (MRI), positron emission tomography (PET), other biological markers, and clinical and neuropsychological assessment can be combined to measure the progression of mild cognitive impairment (MCI) and early Alzheimer’s disease (AD). For up-to-date information, see www.adni-info.org. Demographic, clinical and genomic data have been previously described^30^. Blood-derived mtDNAcn was estimated from WGS as previously described^31^. Linear regression models adjusting for age, sex, *APOE*-ε4 genotypes, diagnosis, and population stratification (PC 1-4) were used to examine the association of mtDNAcn PRS with mtDNAcn.

**Table 2.** Demographic Characteristics of ADGC.

|  | **Alzheimer's Disease** | **Cognitively Unimpaired** |
| --- | --- | --- |
|  | N = 17,005<sup>†</sup> | N = 10,378<sup>†</sup> |
| **Age** | 75.9 ± 8.3 | 73.2 ± 15.8 |
| **Sex** |  |  |
| Female | 10,496 (62%) | 6,112 (59%) |
| Male | 6,509 (38%) | 4,266 (41%) |
| **Ancestry** |  |  |
| AFR | 1,228 (7.2%) | 629 (6.1%) |
| AMR | 1,191 (7.0%) | 674 (6.5%) |
| EAS | 878 (5.2%) | 920 (8.9%) |
| EUR | 13,708 (81%) | 8,155 (79%) |
| **APOE ε4 Carrier** |  |  |
| Non-carrier | 12,539 (74%) | 4,331 (42%) |
| Carrier | 4,466 (26%) | 6,047 (58%) |

<sup>†</sup>Mean ± SD; n (%)

### 2.4 Mendelian Randomization

#### Genetic instrument selection and harmonization of data

To select genetic instruments and harmonize the data, we applied clumping (r² = 0.001, 10Mb window, EUR reference population) to keep independent genome-wide significant SNPs (p < 5e-8). We used LDlinkR version 5 (EUR reference, r² > 0.8) to find LD proxies for instrumental variables absent in the outcome GWAS dataset^32^. The exposure and outcome datasets were harmonized so that SNP effect estimates aligned to the same effect allele, and we used allele frequency information to infer palindromic SNPs^33,34^. We excluded the *APOE* region (19:44912079-19:45912079, build 37) due to its pleiotropic effects^35^.

#### Statistical analysis

MR uses genetic variants as instrumental variables to infer causal relationships between an exposure-outcome pair. Since genetic variants are randomly allocated at conception, they are free from confounding and serve as genetic instruments for causal inference^15^. There are three assumptions in an MR analysis: (1) the genetic variants are associated with the exposure (2) there are no confounders between the genetic variant and outcome (3) all the effects of the genetic variant on the outcome are through the exposure^15^.

Univariate MR was conducted to quantify the causal effect of mtDNAcn on AD and PD using fixed-effects inverse-variance weighted (IVW) approach as the primary analysis method. This method provides a single causal estimate by weighting variant-exposure and variant-outcome associations based on the inverse of their variances, under the assumption that all instruments are valid and there is no horizontal pleiotropy^35^.

#### Diagnostics

We used the Cochran’s Q test to examine heterogeneity and MR Egger regression intercept to detect horizontal pleiotropy. Diagnostics were used to evaluate whether the causal estimates are robust to violations of the MR underlying assumptions^15^. Additionally, we used F-statistics to test the strength of the instruments, whereby higher F-statistics (greater than 10) indicate stronger instruments and reduce the likelihood of weak instrument bias^15^. Outliers were identified and excluded from the MR analysis using Radial MR version 1.0^36^.

#### Sensitivity Analyses

Sensitivity analyses were used to evaluate the robustness of each exposure-outcome analysis under different assumptions, including Random-effects IVW, MR Egger, Weighted Median (WME), and Weighted Mode Based Estimator (WMBE)^37–39^. We defined a robust causal effect as a significant IVW analysis (p<0.05) after outlier removal and where there was no evidence of heterogeneity (p>0.05) or pleiotropy (p>0.05). When there was evidence of heterogeneity or pleiotropy, at least one of the sensitivity analyses also had to be significant (p<0.05) and have the same direction of effect for a robust causal association.

#### Multivariable Mendelian Randomization

MVMR evaluates the direct causal effect of an exposure on an outcome in the presence of other exposures. SNPs are first clumped (r^2^□=□0.001, 10Mb clumping window, EUR reference) and independent genome-wide significant SNPs (p < 5e-8) were extracted from each exposure dataset^15^. For any SNPs not present in either of the exposure datasets, LD proxies are identified and combined. Next, exposure SNPs were extracted from the outcome GWAS, and for SNPs not present in the outcome GWAS, proxy SNPs were identified. TwoSampleMR version 0.5.11 was used to harmonize the mtDNAcn and AD/PD datasets. Finally, MVMR was performed using the MVMR package version 0.3 and MendelianRandomization package version 0.9.0^40,41^.

#### Latent Heritable Confounder Mendelian Randomization

Addressing exchangeability and horizontal pleiotropy is challenging in MR as many SNPs display pleiotropic effects, some of which may be mediated through confounders in the exposure-outcome relationship. LHC-MR addresses these issues by introducing a latent heritable confounder. This unmeasured confounder is a latent factor whose effects are estimated from the pleiotropic effects of instrumental variables (IVs) in the exposure-outcome pairs being analyzed^14^. Including this LHC in the model adjusts for confounding, providing an unbiased beta estimate. Other benefits to LHC-MR is (i) mitigates sample overlap effects by not having to remove overlapping segments which reduces statistical power (ii) enables us to estimate bidirectional effects of the outcome on the exposure (iii) boosts statistical power by using the whole genome to estimate causal effects, rather than just significant SNPs^14^. To correct for multiple testing across the various exposure-outcome pairs analyzed, we applied False Discovery Rate (FDR) correction to the LHC-MR p-values.

The SNPs used as instrumental variables, along with their harmonized effect estimates, are provided in Supplementary Tables 3-14.

## 3. Results

### 3.1 mtDNAcn is negatively correlated with AD and PD

We found strong positive genetic correlations between the four mtDNAcn measures and strong positive genetic correlations between both AD datasets (Figure 1). Longchamps et al was the only mtDNAcn GWAS that had weak negative correlations across AD, AD/dementia, and PD. Gupta et al mtDNAcn was weakly negatively correlated with AD and AD/dementia, while Chong et al mtDNAcn was weakly negatively correlated with AD/dementia. AD/dementia and PD were also weakly negatively correlated (Figure 1).

**Figure 1.**
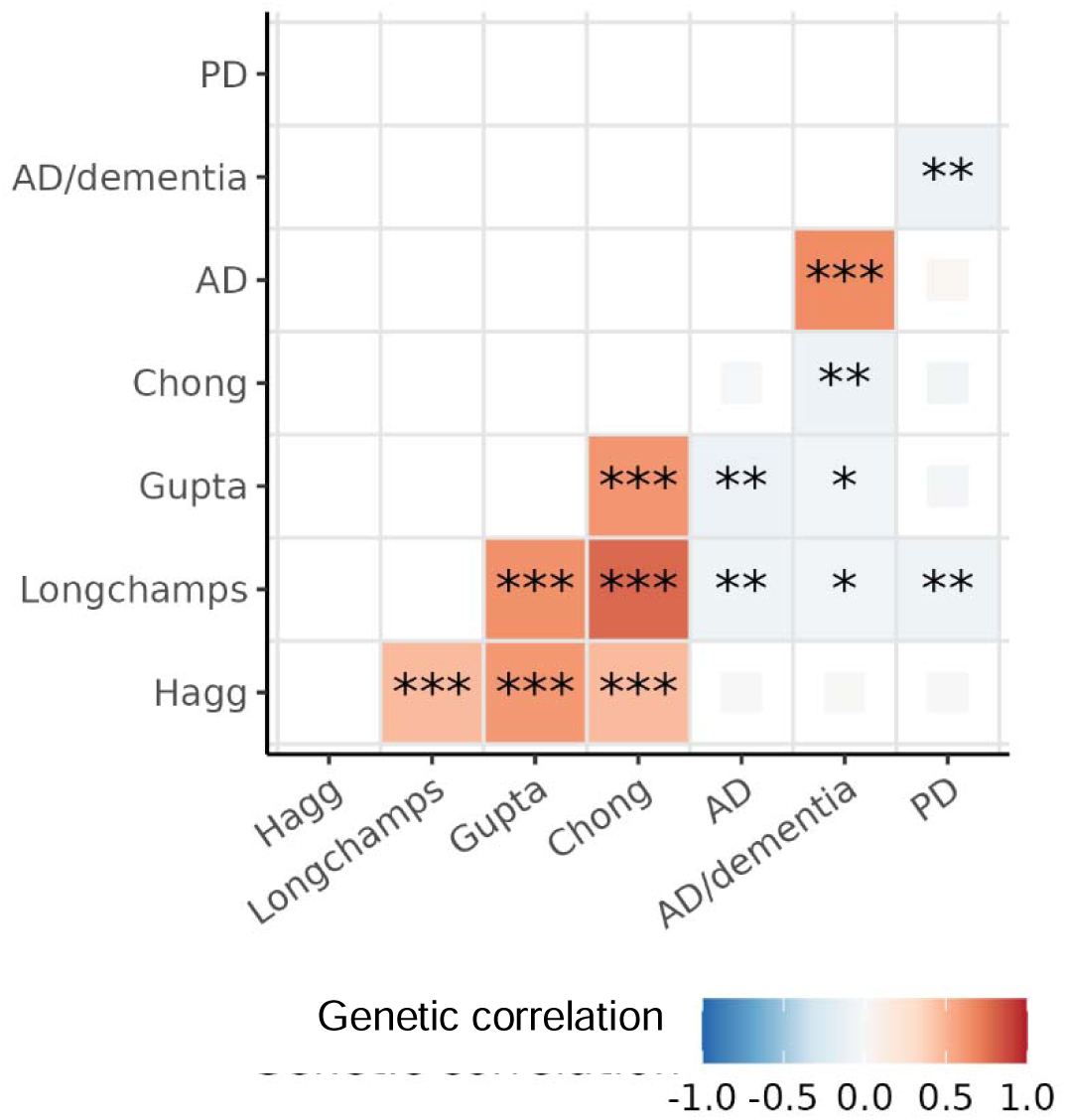
Genetic correlations of mtDNAcn, AD, AD/dementia, and PD. Genetic correlations between four mtDNAcn GWAS, AD, AD/dementia and PD. Box color indicates the direction and magnitude of the genetic correlation (red = positive, blue = negative). Box size reflects statistical significance, with larger tiles indicating significant associations (p ≤ 0.05). Asterisks denote significance thresholds: p ≤ 0.05 (*), p ≤ 0.01 (**), p ≤ 0.001 (***).

### 3.2 mtDNAcn-PRS is not associated with AD

In ADNI, all mtDNAcn PRS models were significantly associated with blood-derived mtDNAcn, with higher genetically predicted mtDNAcn corresponding to higher measured blood-derived mtDNAcn. The Hägg mtDNAcn PRS had the strongest association with mtDNAcn (Supplementary Table 15).

None of the PRS derived from the mtDNAcn GWAS showed a statistically significant association with AD risk in ADGC (Figure 2). However, the Longchamps mtDNAcn PRS showed a trend toward significance (OR [95% CI]= 0.97, [0.95-1.00], p = 0.059).

**Figure 2.**
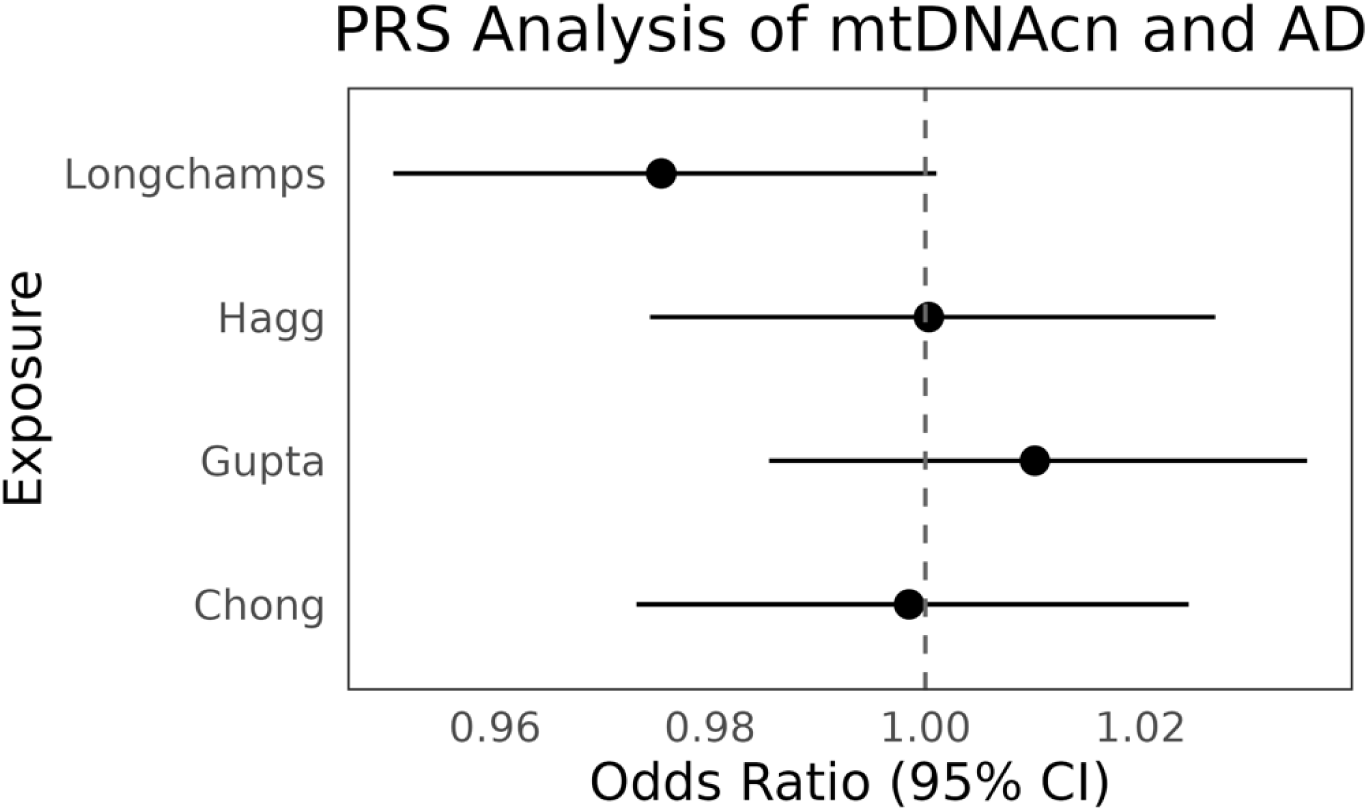
PRS analysis of mtDNAcn and AD. Forest plot showing the PRS odds ratio of four different mtDNAcn measures and AD. Black indicates non-significant results.

### 3.3 Univariable and MVMR showed no causal effect of mtDNAcn on AD or PD

Univariate MR results show no significant causal effect of mtDNAcn onto AD, AD/dementia or PD across all four mtDNAcn measures (Supplementary Table 16, Supplementary Figure 2). While instrument strength was strong in all analyses (F-statistics > 10), significant heterogeneity was observed in nearly all exposure-outcome pairs, with the exception of Gupta et al onto PD (Cochran’s Q value= 33.7, p = 0.33) and Hägg et al onto AD (Cochran’s Q value= 62.2, p = 0.06) (Supplementary Table 16). No pleiotropy was detected in any analysis based on the MR-Egger intercept (Supplementary Table 16). When adjusting for platelets in Hägg using MVMR, we similarly observed no significant causal effect of mtDNAcn on AD or PD (Supplementary Table 17, Supplementary Figure 3). Heterogeneity remained significant, but instruments were strong across all models (Supplementary Table 17).

### 3.4 LHC-MR showed a causal effect of mtDNAcn on AD/dementia and PD, with limited evidence to support a causal association in the reverse direction

LHC-MR results showed that higher genetically predicted mtDNAcn from all four mtDNAcn datasets was significantly associated with lower risk of AD/dementia and PD (Figure 3, Supplementary Table 18). In contrast, higher genetically predicted mtDNAcn from the Longchamps dataset was significantly associated with a higher risk of AD (OR [95% CI]= 1.21, [1.04-1.42], p = 0.01), while the other mtDNAcn datasets yielded non-significant associations with AD. There was significant confounding in all LHC-MR analyses except for Chong onto AD/dementia (p = 0.47) and PD (p = 0.72). Notably, the confounder effect estimates were positive across all AD/dementia and PD analyses, whereas the corresponding causal effect estimates were negative (Supplementary Table 18).

**Figure 3.**
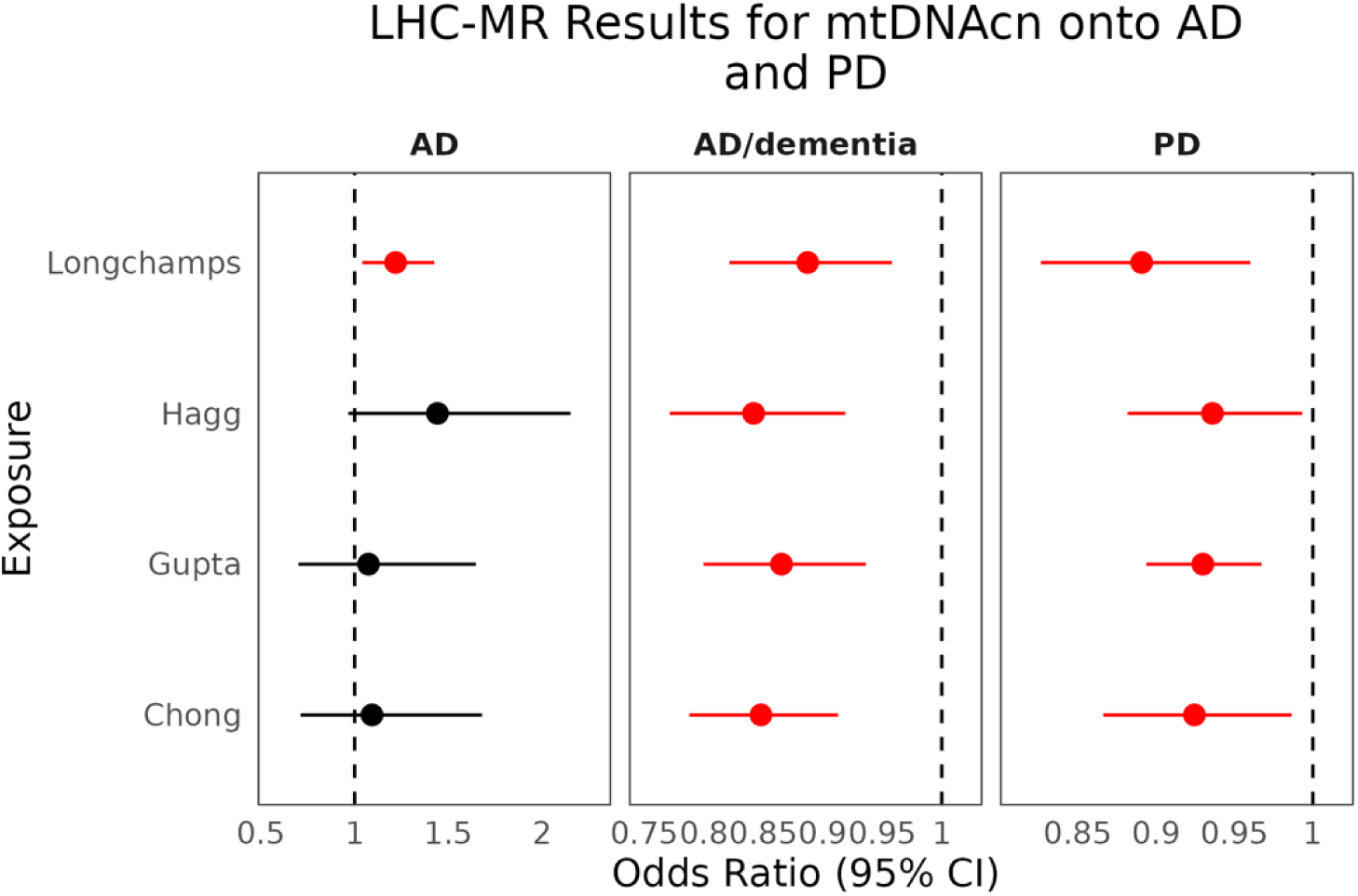
LHC-MR results for mtDNAcn onto AD and PD. Forest plots showing the MR odds ratio for four different mtDNAcn measures, AD, AD/dementia and PD. P values represent FDR-adjusted significance. Red indicates significant results.

In the reverse direction, genetically liability for AD was causally associated with increased mtDNAcn in Longchamps (β [95% CI]= 0.17, [0.08, 0.26], p = 0.002) and Gupta (β [95% CI]= 0.19, [0.07-0.31], p = 0.01)(Supplementary Table 19, Supplementary Figure 4). Furthermore, there was significant confounding for AD onto Longchamps (p=2.91e-25).

## 4. Discussion

This study evaluated the association between genetically predicted mtDNAcn and AD/PD risk across four mtDNAcn GWAS datasets (Longchamps, Hägg, Gupta, Chong). While most analyses yielded consistent results across these datasets, minor discrepancies likely reflect differences in study design, including sample size, platelet contamination, and the method used to measure mtDNAcn. Despite these variations, our findings suggest that different mtDNAcn measurement techniques capture a shared underlying biology. To assess the robustness of these associations, we applied multiple MR methods to evaluate the causal relationships between mtDNAcn measures and both neurodegenerative diseases. The key finding was that higher blood-based mtDNAcn was causally associated with reduced risk of AD/dementia and PD.

Genetic correlations showed that the Longchamps mtDNAcn GWAS was negatively correlated with AD, AD/dementia, and PD, Gupta mtDNAcn GWAS with AD and AD/dementia and Chong mtDNAcn GWAS with AD/dementia. Although none of the PRS associations of mtDNAcn onto AD reached statistical significance, the PRS based on the Longchamps mtDNAcn GWAS showed a trend toward significance. These findings highlight small dataset-specific differences in the strength of association between mtDNAcn genetic risk and AD, which may be explained by how mtDNAcn was quantified. Longchamps was the largest of the four studies, identifying 97 associated loci and estimating mtDNAcn using three different methods: qPCR (1.8% of samples), whole genome sequencing (0.3%), and microarray (97.9%)^19^. Although previous research has shown that mtDNAcn measured by WGS has a stronger correlation with known mtDNA correlates (age, biological sex, white blood cell count, Duffy locus genotype, incident cardiovascular disease), combining all three approaches likely provides the most accurate estimate^3^. Thus, the results from the Longchamps GWAS are likely the most reliable. On the other hand, Hägg et al. did not adjust for platelet counts in their analysis and therefore can be considered the least reliable^20^. Although adjusting for platelets did not change the significance of the MVMR results in our study, prior research has shown that approximately one-third of the genetic associations identified by Hägg et al. lose statistical significance when platelet counts are included as a covariate^21^.

The univariate MR results did not support a causal association between mtDNAcn and AD or PD; however, there was significant heterogeneity in most exposure-outcome pairs, indicating significant violations in the underlying assumptions of MR. In particular, sample overlap between the AD/dementia GWAS and mtDNAcn GWASs—both of which include individuals from UK Biobank—may introduce bias due to the winner’s curse and increased risk of type I errors^42^.

LHC-MR accounts for sample overlap and incorporates a latent, unmeasured confounder —an environmental or genetic factor that influences both the exposure and outcome— and estimates both its presence and effect^14^. LHC-MR results showed that genetically higher mtDNAcn is causally associated with a lower risk of AD/dementia. Additionally, LHC-MR results suggest that mtDNAcn is associated with a lower risk of PD. These results are consistent with prior studies that have found that damaged mtDNA and mtDNA mutations can lead to PD-like pathology^43,44^. Comparing confounder estimates alongside causal estimates helps reveal how confounding may bias causal estimates in univariable MR. In the LHC-MR analysis, heritable confounders showed positive effects, while the causal estimates for mtDNAcn on AD/dementia and PD were negative. These opposing directions may explain why standard MR methods were non-significant.

Higher mtDNAcn in the Longchamps dataset was found to be causally associated with increased AD risk, though this was not replicated for the other mtDNAcn GWASs. These findings may be explained by diagnostic heterogeneity in both AD/dementia and AD GWASs. The AD/dementia GWAS encompasses multiple dementia subtypes, each with distinct underlying risk factors and biological mechanisms^2,45^. Furthermore, the associations observed in the AD/dementia GWAS may also be partly influenced by survival and reporting bias, as many cases are based on self-reported parental diagnoses^46^. However, for the AD GWAS, there may be misclassification in clinical AD, with one study finding that 30% of controls were found to be in preclinical stages of AD even at typical cohort ages^47^. Misassignment can inflate reverse associations, potentially contributing to the reverse causality in our results.

When looking at the bidirectional effects, higher genetic risk of AD was associated with increased mtDNAcn in both the Longchamps and Gupta datasets. This may reflect compensatory upregulation, where mtDNAcn rises in response to functional deficiency regardless of its cause^2^.

There are several limitations in our analyses. First, all GWAS datasets analyzed were predominantly of European ancestry, limiting the generalizability of our findings to other populations. Second, although the Longchamps GWAS included three methods for measuring mtDNAcn, over 98% of samples were measured using microarray therefore it is difficult to draw conclusions about the specific impact of WGS or qPCR. Lastly, while our PRS is based on blood-derived mtDNAcn, it may not fully capture mtDNAcn in brain tissue where AD manifests, and baseline blood-based mtDNAcn levels may not reflect the brain’s adaptive capacity to compensate for age-related mitochondrial decline.

Strengths of our study include the use of multiple mtDNAcn and AD GWAS that were conducted using different study designs. Different mtDNAcn GWASs rely on distinct participant samples and analytical approaches, while the AD GWASs rely on different definitions of cases versus controls based on underlying AD pathology. These differences can affect the estimated associations between SNPs and the trait and introduce variability that can impact MR causal effect estimates^48^. Another strength is a bidirectional MR approach to assess the directionality of effects and the application of both PRS and MR to distinguish association from causation.

In conclusion, our study demonstrates a causal effect of mtDNAcn on AD and PD using multiple complementary analytical approaches and different mtDNAcn measures. Additional research is needed to extend these findings to other AD endophenotypes to better understand the biological pathways underlying these findings. Nevertheless, these findings highlight mtDNAcn as an important biomarker of mitochondrial dysfunction in the early stages of AD and PD.

## Supporting information

STROBE-MR Checklist

STables 1-2; SFigures 1-4

STables 3-14

STables 15-19

## Data Availability

All statistical analyses were conducted using R (v4.3.0). The analysis code is publicly available at: https://github.com/AndrewsLabUCSF/Brian-Estimating-Causality-mtDNAcn-on-Alzheimer/tree/aadrita_mtdnacn. The original summary statistics are available at the following websites: https://dss.niagads.org/datasets/ng00075/,

https://www.ebi.ac.uk/gwas/publications/35379992,

https://pmc.ncbi.nlm.nih.gov/articles/PMC8422160/#S15,

https://www.ebi.ac.uk/gwas/publications/27863252,

https://www.ebi.ac.uk/gwas/publications/35023831,

https://link.springer.com/article/10.1007/s00439-020-02249-w#data-availability,

https://pmc.ncbi.nlm.nih.gov/articles/PMC8758627/#notes2,

https://www.ebi.ac.uk/gwas/publications/37587338

## Acknowledgements

ADNI

**Data used in preparation of this article were obtained from the Alzheimer’s Disease Neuroimaging Initiative (ADNI) database (adni.loni.usc.edu). As such, the investigators within the ADNI contributed to the design and implementation of ADNI and/or provided data but did not participate in analysis or writing of this report. A complete listing of ADNI investigators can be found at: http://adni.loni.usc.edu/wp-content/uploads/how_to_apply/ADNI_Acknowledgement_List.pdf

Data collection and sharing for this project was funded by the Alzheimer’s Disease Neuroimaging Initiative (ADNI) (National Institutes of Health Grant U01 AG024904) and DOD ADNI (Department of Defense award number W81XWH-12-2-0012). ADNI is funded by the National Institute on Aging, the National Institute of Biomedical Imaging and Bioengineering, and through generous contributions from the following: AbbVie, Alzheimer’s Association; Alzheimer’s Drug Discovery Foundation; Araclon Biotech; BioClinica, Inc.; Biogen; Bristol-Myers Squibb Company; CereSpir, Inc.; Cogstate; Eisai Inc.; Elan Pharmaceuticals, Inc.; Eli Lilly and Company; EuroImmun; F. Hoffmann-La Roche Ltd and its affiliated company Genentech, Inc.; Fujirebio; GE Healthcare; IXICO Ltd.; Janssen Alzheimer Immunotherapy Research & Development, LLC.; Johnson & Johnson Pharmaceutical Research & Development LLC.; Lumosity; Lundbeck; Merck & Co., Inc.; Meso Scale Diagnostics, LLC.; NeuroRx Research; Neurotrack Technologies; Novartis Pharmaceuticals Corporation; Pfizer Inc.; Piramal Imaging; Servier; Takeda Pharmaceutical Company; and Transition Therapeutics. The Canadian Institutes of Health Research is providing funds to support ADNI clinical sites in Canada. Private sector contributions are facilitated by the Foundation for the National Institutes of Health (www.fnih.org). The grantee organization is the Northern California Institute for Research and Education, and the study is coordinated by the Alzheimer’s Therapeutic Research Institute at the University of Southern California. ADNI data are disseminated by the Laboratory for Neuro Imaging at the University of Southern California.

---

**Supplementary Table 1. Description of GWAS datasets used in this study**

**Supplementary Table 2. Demographic Characteristics of ADNI**

**Supplementary Table 3: Harmonized data for Chong mtdnacn on AD univariate MR analysis**

**Supplementary Table 4: Harmonized data for Chong mtdnacn on AD/dementia univariate MR analysis**

**Supplementary Table 5: Harmonized data for Chong mtdnacn on PD univariate MR analysis**

**Supplementary Table 6: Harmonized data for Longchamps mtdnacn on AD univariate MR analysis**

**Supplementary Table 7: Harmonized data for Longchamps mtdnacn on AD/dementia univariate MR analysis**

**Supplementary Table 8: Harmonized data for Longchamps mtdnacn on PD univariate MR analysis**

**Supplementary Table 9: Harmonized data for Gupta mtdnacn on AD univariate MR analysis**

**Supplementary Table 10: Harmonized data for Gupta mtdnacn on AD/dementia univariate MR analysis**

**Supplementary Table 11: Harmonized data for Gupta mtdnacn on PD univariate MR analysis**

**Supplementary Table 12: Harmonized data for Hägg mtdnacn on AD univariate MR analysis**

**Supplementary Table 13: Harmonized data for Hägg mtdnacn on AD/dementia univariate MR analysis**

**Supplementary Table 14: Harmonized data for Hägg mtdnacn on PD univariate MR analysis**

**Supplementary Table 15: PRS associations of mtDNAcn on mtDNAcn in ADNI**

**Supplementary Table 16. Summary of univariable MR results with AD, AD/dementia and PD**

**Supplementary Table 17. Summary of multivariable MR results of Hägg onto AD, AD/dementia and PD adjusting for platelets**

**Supplementary Table 18: LHC-MR results forward direction**

**Supplementary Table 19: LHC-MR results reverse direction**

**Supplementary Fig 1. Overview of the study design**

**Supplementary Fig 2. Univariate MR results for mtDNAcn onto AD and PD**

**Supplementary Fig 3. MVMR odds ratio for Hägg**

**Supplementary Fig 4. LHC-MR results for AD and PD onto mtDNAcn STROBE MR Checklist**

