## Supplementary material for "Evaluating the causal effect of mitochondrial dysfunction on Alzheimer’s and Parkinson’s disease using Polygenic Risk Scores and Mendelian Randomization": STables 1-2; SFigures 1-4

**Table of Contents**

Supplementary Table S1. Description of GWAS datasets used in this study	4

Supplementary Table S2. Demographic Characteristics of ADNI	6

Supplementary Fig 1. Overview of the study design	7

Supplementary Fig 2. Univariate MR results for mtDNAcn onto AD and PD	8

Supplementary Fig 3. MVMR odds ratio for Hägg	9

Supplementary Fig 4. LHC-MR results for AD and PD onto mtDNAcn	10

**Supplementary Table S1. Description of GWAS datasets used in this study**

| Study | Trait | Details | Mean age |
| --- | --- | --- | --- |
| Kunkle et al, 2019 | Late-onset Alzheimer’s disease | The study was conducted in three stages. Stage 1 was a meta-analysis of non-Hispanic White (NHW) individuals across 46 datasets, including 21,982 Alzheimer’s disease (AD) cases and 41,944 cognitively normal controls. Stage 2 involved replication using a custom genotyping array. Stage 3 was divided into two sub-cohorts: Stage 3A (n = 11,666) and Stage 3B (n = 30,511). AD cases were defined either by clinical diagnosis or confirmed by autopsy. In total, the study included 35,274 AD cases and 59,163 controls. | 58-86 |
| Bellenguez et al, 2022 | AD/dementia | The study was conducted in two stages. Stage 1 included the European Alzheimer & Dementia Biobank (EADB), which aggregated data from multiple European GWAS consortia, and was meta-analyzed with proxy-AD data from the UK Biobank (UKB). This stage comprised 39,106 clinically diagnosed AD cases, 46,828 proxy cases from UKB, and 401,577 controls, following standard quality control procedures. Stage 2 included data from the ADGC, FinnGen, and CHARGE consortia, totaling 25,392 AD cases and 276,086 controls. | N/A |
| Astle et al, 2016 | Platelets | This study conducted analyses using data from three large cohorts with available blood cell trait measurements and genome-wide imputed genetic data. Quality control measures were applied at multiple stages, including array-specific filters for call rate, Hardy-Weinberg equilibrium, plate and batch effects, heterozygosity outliers, and ancestry outliers determined by principal components analysis. Additional quality control steps removed duplicate samples, non-European ancestry individuals, and samples with excessive contamination. | N/A |
| Chong M et al, 2022 | Mitochondrial copy number | A novel array-based method for estimating mitochondrial DNA copy number (mtDNAcn), called Automatic Mitochondrial Copy (AutoMitoC), was developed and applied to study participants. UK Biobank genetic data was used to develop the AutoMitoC pipeline and validated in the INTERSTROKE study. After standard quality control procedures—including exclusions based on sex mismatch, genotype call rate, and heterozygosity—genotype imputation was performed using the Haplotype Reference Consortium and UK10K panels. Genetic variants were filtered using thresholds for imputation quality and minor allele frequency. Ethical approval and informed consent were obtained as part of UK Biobank’s centralized protocol. | 56.9 |
| Hägg S et al 2021 | Mitochondrial copy number | This study used data from the UKB. Individuals with low call rates, high heterozygosity, sex discordance, relatedness, or non-European ancestry were removed. Mitochondrial DNA (mtDNA) abundance was estimated using array-based genotyping probe intensities on the mitochondrial chromosome. A novel weighted approach, derived from a multivariate regression against exome-based mtDNA coverage, was developed to more accurately reflect true mtDNA abundance. This estimate was standardized within genotyping plates and validated against known biological correlates such as age, sex, BMI, smoking, and blood cell counts. The GWAS of mtDNA abundance was performed using PLINK2, adjusting for age, sex, genotyping batch, call rate, white blood cell metrics, and population structure. Variants included in the GWAS passed strict filters for call rate and imputation quality. All participants provided informed consent, and the study was approved under UKB project ID 22,224. | 56.6 |
| LongChamps et al, 2021 | Mitochondrial copy number | MtDNAcn was estimated using three main approaches: quantitative PCR (qPCR), microarray-based intensity methods across several cohorts using the Genvisis MitoPipeline, and whole-genome sequencing. The UKB cohort used a hybrid method that leveraged both whole-exome sequencing and genotyping array data, adjusted for technical artifacts and blood cell counts. Covariates such as age, sex, and blood cell composition were adjusted for in all cohorts, and standardized residuals of mtDNA-CN were used for analysis. Genome-wide association analyses were performed using linear regression within each cohort, followed by ancestry-stratified meta-analyses using a random-effects model. Participants provided informed consent and all protocols received IRB approval. | 56.6 |
| Gupta et al, 2023 | Mitochondrial copy number | This study used mtSwirl, a scalable pipeline for mtDNAcn estimation and variant calling. The pipeline integrates whole-genome sequencing (WGS) data and supports single- or multi-sample execution. mtSwirl was applied to four major cohorts: the UKB, All of Us (AoU), gnomAD v3.1, and the 1000 Genomes Project (1000G). UKB and AoU samples were derived from blood and sequenced using harmonized protocols, while gnomAD and 1000G included diverse global populations with high-coverage WGS data, excluding cell lines and low-quality samples. MtDNAcn was computed using read depth ratios, and variant and sample quality control included thresholds for contamination, heteroplasmy, and sequencing artifacts. | 20-90 in AoU and 40-70 for UKB |
| Nalls et al, 2019 | Parkinson’s Disease | This genome-wide association study (GWAS) of Parkinson’s disease (PD) leveraged three primary sources of data for discovery analyses: (1) previously published datasets (2) 13 newly curated case-control datasets, and (3) UKB proxy-case data. Across all sources, uniform quality control procedures were applied, including exclusion of related individuals, non-European ancestry samples, and SNPs with low call rate, MAF <1%, or poor imputation quality. The UKB data utilized a genome-wide association by proxy (GWAX) approach, with proxy cases defined by reported first-degree relatives with PD. Diagnostic criteria for PD included clinical diagnosis aligned with UK Brain Bank criteria or self-report, depending on the cohort. A total of 37,688 cases, 18,618 proxy cases, and 1,474,097 controls contributed to a fixed-effects meta-analysis across 7.8 million SNPs. Genotype imputation was performed using 1000 Genomes or the HRC reference panel depending on the dataset. All studies received ethics committee approval, and participants provided informed consent for genetic research. | N/A |

**Supplementary Table S2. Demographic Characteristics of ADNI**

| **Variable** | **Cognitively Normal**, N = 300*^1^* | **Significant Memory Concern**, N = 99*^1^* | **Mild Cognitive Impairment**, N = 710*^1^* | **Alzheimer’s Disease**, N = 246*^1^* |
| --- | --- | --- | --- | --- |
| **Age** | 75.0 ± 5.5 | 72.3 ± 5.6 | 73.0 ± 7.5 | 74.9 ± 7.9 |
| **Sex** |  |  |  |  |
| Female | 141 (47%) | 58 (59%) | 283 (40%) | 105 (43%) |
| Male | 159 (53%) | 41 (41%) | 427 (60%) | 141 (57%) |
| **Race** |  |  |  |  |
| Non-White | 4 (1.3%) | 1 (1.0%) | 7 (1.0%) | 1 (0.4%) |
| White | 296 (99%) | 98 (99%) | 703 (99%) | 245 (100%) |
| **APOE ε4 Carrier Status** |  |  |  |  |
| Carrier | 80 (27%) | 34 (34%) | 355 (50%) | 166 (67%) |
| Non-carrier | 220 (73%) | 65 (66%) | 355 (50%) | 80 (33%) |
| **mtcn** | 161.6 ± 33.3 | 226.9 ± 47.4 | 171.8 ± 47.1 | 178.6 ± 42.8 |
| *^1^*Mean ± SD; n(%) |  |  |  |  |

**Supplementary Fig 1. Overview of study design**

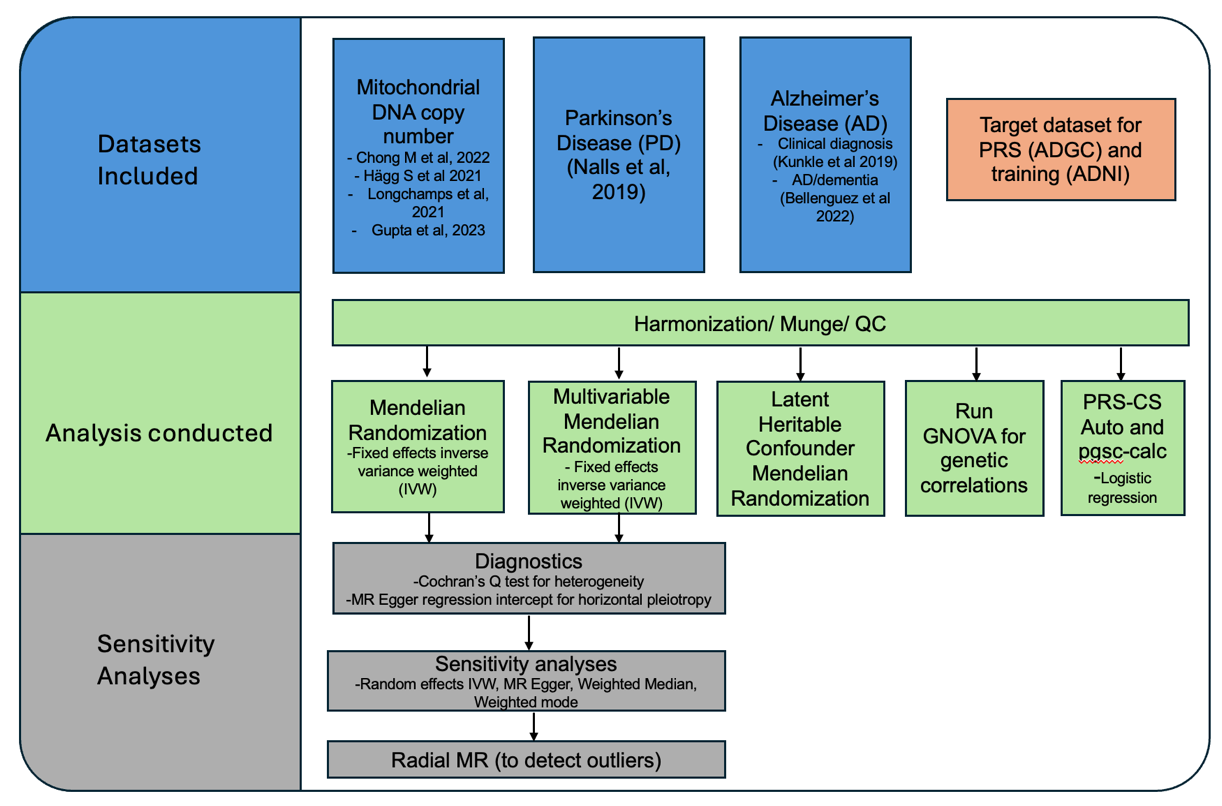

In ADGC, ancestry-normalized PRS were generated using all four mtDNAcn GWAS (PRS-CS-auto, 1000 Genomes European reference panel, HapMap3 SNPs, excluding *APOE* region). Logistic regression assessed the association between mtDNAcn PRS and AD status, adjusting for age, sex, *APOE*-ε4 status and population stratification. For MR, Inverse Variance Weighted was the primary method with sensitivity analyses. MVMR was used to adjust mtDNAcn for the effects of platelets for the Hägg GWAS and LHC-MR was used to estimate the causal effect of mtDNAcn on AD while accounting for unmeasured confounding.

**Supplementary Fig 2. Univariate MR results for mtDNAcn onto AD and PD**

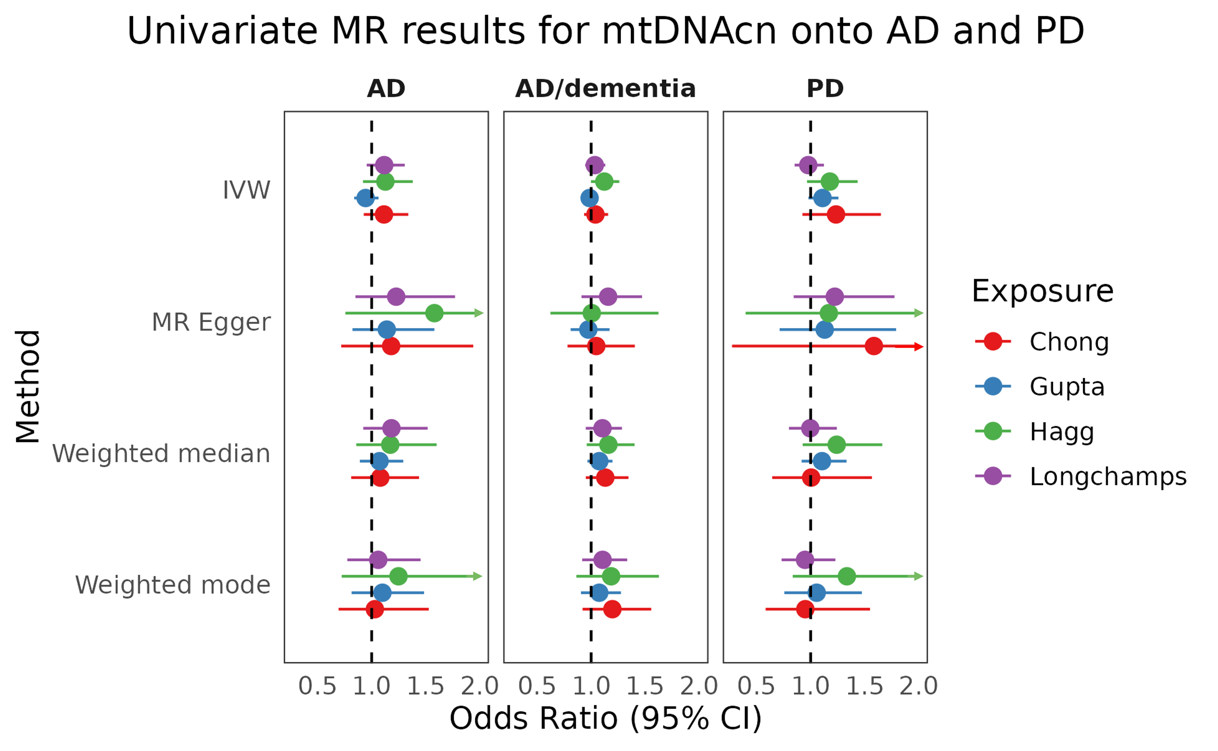

Forest plots showing the univariable MR odds ratio for the four measures of mtDNAcn onto AD, AD/dementia and PD. Arrows represent CI intervals that continue past the x axis.

**Supplementary Fig 3. MVMR odds ratio for Hägg**

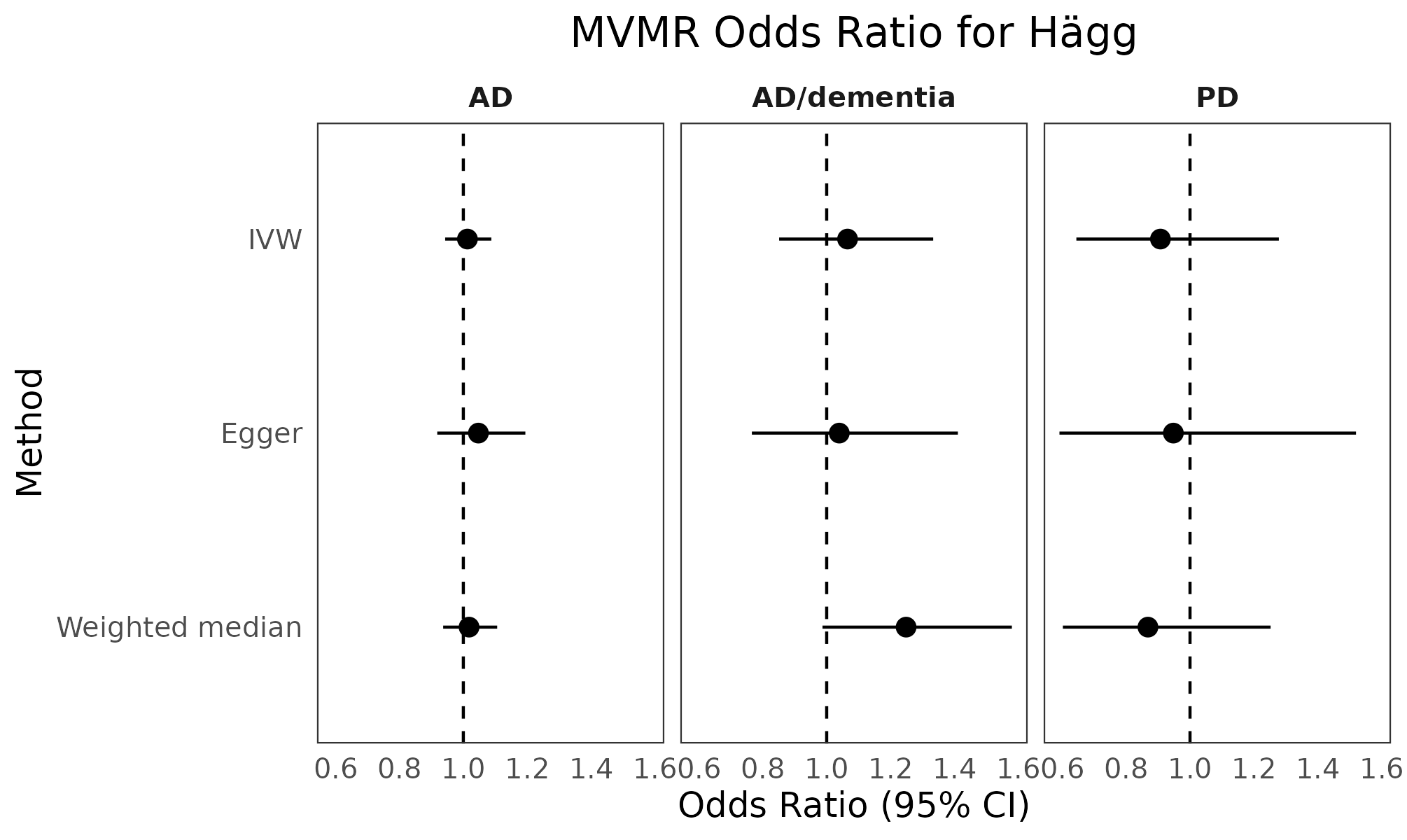

Forest plots showing the multivariable MR odds ratio for Hägg mtDNAcn GWAS after adjusting for platelets. Black indicates non-significant results.

**Supplementary Fig 4. LHC-MR results for AD and PD onto mtDNAcn**

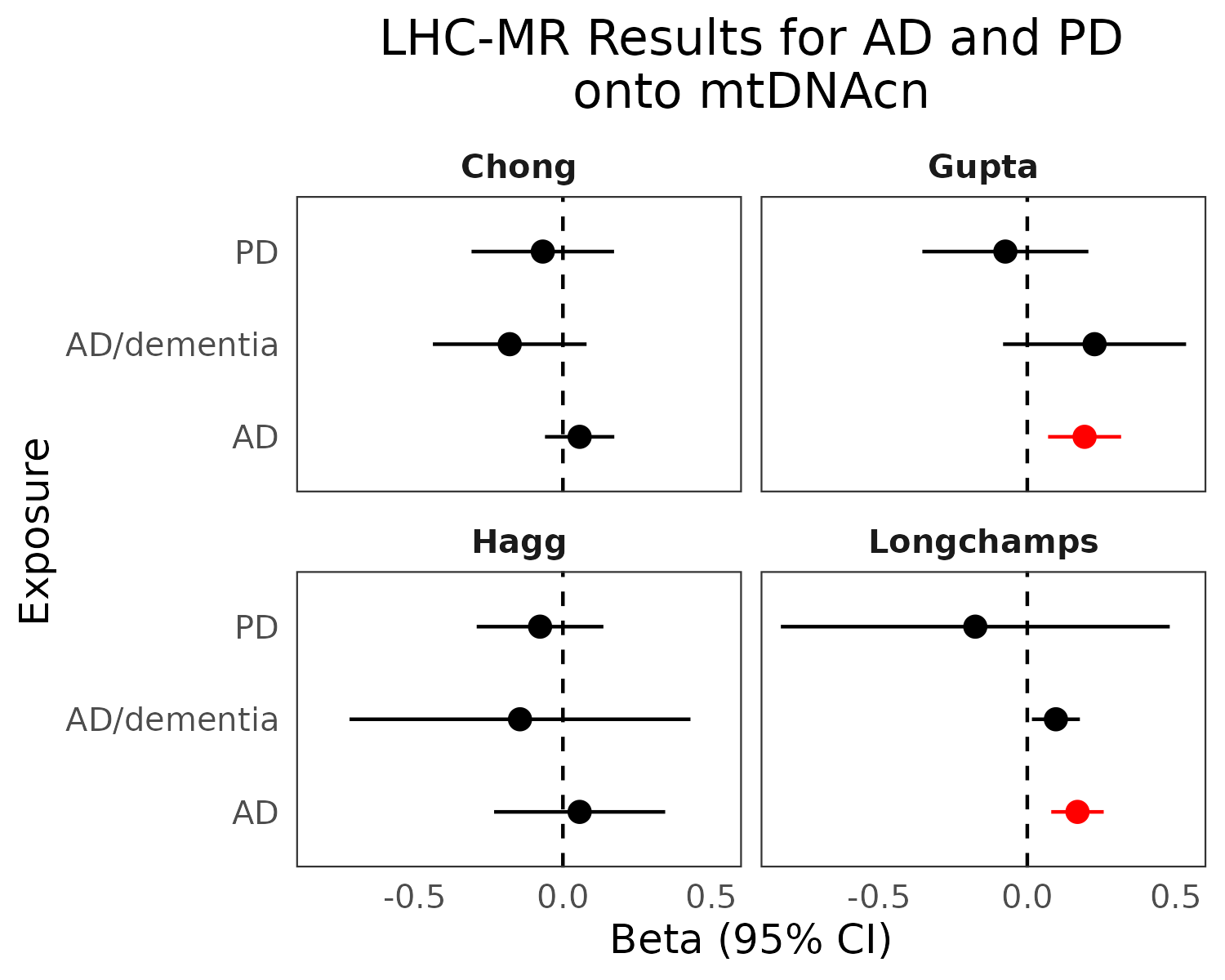

Forest plots showing the MR odds ratio for AD, AD/dementia and PD and four different measures of mtDNAcn. P values represent FDR-adjusted significance. Red indicates significant results.
