## Supplementary material for "Evaluating the causal effect of mitochondrial dysfunction on Alzheimer’s and Parkinson’s disease using Polygenic Risk Scores and Mendelian Randomization": STables 15-19

^9^ Alzheimer’s Disease Research Center, Keck School of Medicine, University of Southern California, Los Angeles, CA, USA

^10^ Laboratory of Neuroimaging, USC Stevens Neuroimaging and Informatics Institute, Keck School of Medicine, University of Southern California, Los Angeles, CA, USA

**Table of Contents**

Supplementary Table S15: PRS associations of mtDNAcn on mtDNAcn in ADNI	4

Supplementary Table S16. Summary of univariable MR results with AD, AD/dementia and PD	5

Supplementary Table S17. Summary of multivariable MR results of Hägg onto AD, AD/dementia and PD adjusting for platelets. 6

Supplementary Table S18. LHC-MR results forward direction	7

Supplementary Table S19. LHC-MR results reverse direction	8

**Supplementary Table S15: PRS associations of mtDNAcn on mtDNAcn in ADNI**

| Exposure | Outcome | Estimate | SE | P-value |
| --- | --- | --- | --- | --- |
| Chong | mtDNAcn | 4.88 | 1.17 | 3.58E-05 |
| Longchamps | mtDNAcn | 5.33 | 1.15 | 4.64E-06 |
| Hagg | mtDNAcn | 5.14 | 1.22 | 3.00E-05 |
| Gupta | mtDNAcn | 4.86 | 1.09 | 1.16E-05 |

**Supplementary Table S16. Summary of univariable MR results with AD, AD/dementia and PD**

|  |  | SNP | F stats |  | Fixed effects IVW | |  | MR-Egger | |  | WME | |  | WMBE | |  | Cochran’s Q test | MR-Egger intercept |
| --- | --- | --- | --- | --- | --- | --- | --- | --- | --- | --- | --- | --- | --- | --- | --- | --- | --- | --- |
| Exposure | Outcome |  |  |  | Beta (se) | P value |  | Beta (se) | P value |  | Beta (se) | P value |  | Beta (se) | P value |  | Q p value | P value |
| Chong | AD | 62 | 92.9 |  | 0.11 (0.09) | 0.250 |  | 0.17 (0.25) | 0.516 |  | 0.08 (0.15) | 0.601 |  | 0.03 (0.2) | 0.886 |  | 0.02 | 0.80 |
| Chong | AD/dementia | 59 | 92.9 |  | 0.04 (0.05) | 0.474 |  | 0.05 (0.15) | 0.758 |  | 0.12 (0.09) | 0.166 |  | 0.18 (0.13) | 0.186 |  | 0.006 | 0.95 |
| Chong | PD | 14 | 69.3 |  | 0.21 (0.15) | 0.154 |  | 0.46 (0.9) | 0.617 |  | 0 (0.23) | 0.983 |  | -0.05 (0.25) | 0.842 |  | 0.001 | 0.77 |
| Gupta | AD | 63 | 75.12 |  | -0.06 (0.06) | 0.342 |  | 0.13 (0.17) | 0.435 |  | 0.07 (0.09) | 0.459 |  | 0.09 (0.15) | 0.539 |  | 0.019 | 0.21 |
| Gupta | AD/dementia | 61 | 74.6 |  | -0.01 (0.03) | 0.675 |  | -0.03 (0.09) | 0.777 |  | 0.07 (0.05) | 0.187 |  | 0.07 (0.09) | 0.413 |  | 0.019 | 0.88 |
| Gupta | PD | 33 | 58.12 |  | 0.1 (0.06) | 0.101 |  | 0.12 (0.23) | 0.606 |  | 0.1 (0.1) | 0.296 |  | 0.05 (0.17) | 0.752 |  | 0.33 | 0.93 |
| Hägg | AD | 48 | 64.07 |  | 0.12 (0.1) | 0.249 |  | 0.46 (0.38) | 0.230 |  | 0.16 (0.16) | 0.320 |  | 0.22 (0.28) | 0.431 |  | 0.06 | 0.34 |
| Hägg | AD/dementia | 47 | 63.1 |  | 0.11 (0.06) | 0.054 |  | 0.01 (0.24) | 0.981 |  | 0.15 (0.1) | 0.127 |  | 0.17 (0.16) | 0.300 |  | 0.002 | 0.63 |
| Hägg | PD | 26 | 54.7 |  | 0.16 (0.1) | 0.105 |  | 0.16 (0.55) | 0.779 |  | 0.22 (0.15) | 0.148 |  | 0.29 (0.24) | 0.239 |  | 0.013 | 0.98 |
| Longchamps | AD | 89 | 89.4 |  | 0.11 (0.08) | 0.170 |  | 0.2 (0.19) | 0.279 |  | 0.17 (0.13) | 0.186 |  | 0.06 (0.16) | 0.714 |  | 0.017 | 0.56 |
| Longchamps | AD/dementia | 88 | 88.7 |  | 0.03 (0.05) | 0.472 |  | 0.15 (0.12) | 0.236 |  | 0.1 (0.08) | 0.196 |  | 0.1 (0.1) | 0.294 |  | 8.6e-06 | 0.28 |
| Longchamps | PD | 57 | 81.1 |  | -0.02 (0.07) | 0.752 |  | 0.2 (0.19) | 0.293 |  | 0 (0.11) | 0.975 |  | -0.05 (0.13) | 0.689 |  | 0.045 | 0.19 |

**Supplementary Table S17. Summary of multivariable MR results of Hägg onto AD, AD/dementia and PD adjusting for platelets**

|  | | SNP | F stats |  | Fixed- effects IVW | |  | MR-Egger | |  | Weighted median | |  |  | | Cochran’s Q test | |
| --- | --- | --- | --- | --- | --- | --- | --- | --- | --- | --- | --- | --- | --- | --- | --- | --- | --- |
| Exposure | Outcome |  |  |  | Beta (se) | P value |  | Beta (se) | P value |  | Beta (se) | P value |  | |  | | Q p value |
| Platelets | AD | 89 | 43.2 |  | 0.05 (0.1) | 0.60 |  | 0.05 (0.1) | 0.56 |  | -0.09 (0.12) | 0.43 |  | |  | | 4.93e-10 |
| Hägg | AD | 29 | 19.5 |  | 0.01 (0.03) | 0.72 |  | 0.04 (0.06) | 0.49 |  | 0.01 (0.04) | 0.67 |  | |  | |  |
| Platelets | AD/dementia | 89 | 43.2 |  | -0.007 (0.04) | 0.85 |  | -0.008 (0.04) | 0.84 |  | -0.02 (0.03) | 0.49 |  | |  | | 1.37e-20 |
| Hägg | AD/dementia | 29 | 19.5 |  | 0.06 (0.11) | 0.58 |  | 0.03 (0.15) | 0.80 |  | 0.22 (0.12) | 0.06 |  | |  | |  |
| Platelets | PD | 10 | 11.3 |  | -0.017 (0.04) | 0.72 |  | -0.014 (0.05) | 0.77 |  | -0.003 (0.05) | 0.94 |  | |  | | 0.0002 |
| Hägg | PD | 64 | 36.01 |  | -0.09 (0.17) | 0.57 |  | -0.05 (0.24) | 0.82 |  | -0.14 (0.18) | 0.45 |  | |  | |  |

**Supplementary Table S18. LHC-MR results forward direction**

| Exposure | Outcome | Confounder Beta | Confounder SE | Confounder Pvalue | Beta | SE | OR | OR_lci95 | OR_uci95 | Pval | Pval_fdr |
| --- | --- | --- | --- | --- | --- | --- | --- | --- | --- | --- | --- |
| Chong | AD | 0.06130 | 0.02470 | 0.01307 | 0.08910 | 0.21910 | 1.09319 | 0.71154 | 1.67957 | 0.68424 | 0.74249 |
| Longchamps | AD | 0.09355 | 0.01085 | 0.00000 | 0.19778 | 0.08008 | 1.21869 | 1.04166 | 1.42581 | 0.01352 | 0.02318 |
| Hagg | AD | 0.07571 | 0.01729 | 0.00001 | 0.36633 | 0.20409 | 1.44243 | 0.96688 | 2.15187 | 0.07266 | 0.08719 |
| Gupta | AD | 0.10322 | 0.02867 | 0.00032 | 0.07168 | 0.21817 | 1.07431 | 0.70052 | 1.64755 | 0.74249 | 0.74249 |
| Chong | AD/dementia | 0.01677 | 0.02344 | 0.47447 | -0.16459 | 0.03752 | 0.84824 | 0.78810 | 0.91298 | 0.00001 | 0.00014 |
| Longchamps | AD/dementia | 0.05600 | 0.02105 | 0.00779 | -0.11945 | 0.03918 | 0.88741 | 0.82181 | 0.95825 | 0.00230 | 0.00495 |
| Hagg | AD/dementia | 0.06425 | 0.02303 | 0.00527 | -0.17190 | 0.04463 | 0.84206 | 0.77154 | 0.91903 | 0.00012 | 0.00070 |
| Gupta | AD/dementia | 0.08205 | 0.03458 | 0.01765 | -0.14446 | 0.04010 | 0.86549 | 0.80007 | 0.93627 | 0.00032 | 0.00095 |
| Chong | PD | 0.00881 | 0.02461 | 0.72037 | -0.07847 | 0.03310 | 0.92453 | 0.86645 | 0.98650 | 0.01776 | 0.02663 |
| Longchamps | PD | 0.04770 | 0.02342 | 0.04168 | -0.11549 | 0.03817 | 0.89093 | 0.82671 | 0.96013 | 0.00248 | 0.00495 |
| Hagg | PD | 0.08666 | 0.01811 | 0.00000 | -0.06611 | 0.03026 | 0.93603 | 0.88213 | 0.99323 | 0.02892 | 0.03855 |
| Gupta | PD | 0.09862 | 0.02894 | 0.00066 | -0.07265 | 0.02011 | 0.92992 | 0.89398 | 0.96731 | 0.00030 | 0.00095 |

**Supplementary Table S19. LHC-MR results reverse direction**

| Exposure | Outcome | Confounder Beta | Confounder SE | Confounder Pvalue | beta | SE | CI_lci95 | CI_uci95 | Pval | Pval_fdr |
| --- | --- | --- | --- | --- | --- | --- | --- | --- | --- | --- |
| AD | Chong | -0.16579 | 0.05470 | 0.00244 | 0.05644 | 0.05959 | -0.06035 | 0.17323 | 0.34353 | 0.67831 |
| AD | Longchamps | -0.26158 | 0.02519 | 0.00000 | 0.16911 | 0.04510 | 0.08071 | 0.25751 | 0.00018 | 0.00213 |
| AD | Hagg | -0.12756 | 0.03228 | 0.00008 | 0.05644 | 0.14709 | -0.23185 | 0.34472 | 0.70121 | 0.70121 |
| AD | Gupta | -0.13964 | 0.08071 | 0.08360 | 0.19310 | 0.06276 | 0.07009 | 0.31610 | 0.00209 | 0.01255 |
| AD/dementia | Chong | -0.03676 | 0.02208 | 0.09601 | -0.17915 | 0.13212 | -0.43810 | 0.07980 | 0.17509 | 0.42022 |
| AD/dementia | Longchamps | -0.04383 | 0.01516 | 0.00383 | 0.09639 | 0.04121 | 0.01562 | 0.17717 | 0.01934 | 0.07737 |
| AD/dementia | Hagg | 0.04108 | 0.07707 | 0.59402 | -0.14460 | 0.29311 | -0.71910 | 0.42991 | 0.62179 | 0.67831 |
| AD/dementia | Gupta | 0.05443 | 0.01740 | 0.00175 | 0.22672 | 0.15735 | -0.08167 | 0.53512 | 0.14961 | 0.42022 |
| PD | Chong | -0.02613 | 0.01300 | 0.04440 | -0.06774 | 0.12250 | -0.30785 | 0.17237 | 0.58030 | 0.67831 |
| PD | Longchamps | 0.01586 | 0.03675 | 0.66615 | -0.17523 | 0.33432 | -0.83049 | 0.48004 | 0.60019 | 0.67831 |
| PD | Hagg | 0.01287 | 0.03106 | 0.67856 | -0.07697 | 0.10892 | -0.29044 | 0.13650 | 0.47975 | 0.67831 |
| PD | Gupta | 0.03069 | 0.01077 | 0.00437 | -0.07392 | 0.14273 | -0.35367 | 0.20584 | 0.60453 | 0.67831 |
